# Estimating malaria attributable fraction using quantitative PCR in a longitudinal cohort in Eastern Uganda

**DOI:** 10.64898/2026.02.24.26347052

**Authors:** Anne C Martin, Qiyu Wang, Shakira Babirye, Emmanuel Arinaitwe, Maato Zedi, Isaac Ssewanyana, Felistas Nankya Namirimu, Patience Nayebare, Peter Olwoch, Stephen Tukwasibwe, Prasanna Jagannathan, Joaniter I. Nankabirwa, Moses Kamya, Grant Dorsey, Bryan Greenhouse, Jessica Briggs, Isabel Rodriguez-Barraquer

## Abstract

Persistent, asymptomatic *Plasmodium* infections are common in areas of high transmission due to acquired immunity. When asymptomatically infected individuals seek care for a fever caused by something other than malaria parasites, they may test positive for parasites and be incorrectly diagnosed as having clinical malaria. This study used distributions of qPCR parasite densities to estimate the fraction of fever attributable to malaria (malaria attributable fraction, MAF) in a cohort of 659 individuals followed for up to 3 years from three geographically distinct zones in eastern Uganda. Prevalence of *P. falciparum* by qPCR ranged from 47-63% in the three zones, with over 95% of cohort members parasitemic at least once. Overall MAF across the three zones ranged from 54-64%. MAF was highest in under-five year-olds (72%), next highest in 5-15 year-olds (56%) and lowest in adults over 16 (45%). Notably, nearly 50% of fevers with low to moderate parasite density (10 - 100 parasite/ microliter) were attributed to malaria. MAF-corrected incidence was higher than the definition of clinical malaria used in many vaccine field-studies (fever and parasite density ≥ 5000/microliter) and the difference varied by age group: MAF-corrected incidence was 18% higher in children under five, 7% higher in 5-15 year olds, and 70% higher in adults. These results suggest parasite density thresholds commonly used to define primary study outcomes will underestimate the true incidence of clinical malaria. Studies aiming to precisely estimate intervention effects on incidence should consider estimating MAF in their study population and incorporating it into their design.

## Introduction

Clinical malaria due to *Plasmodium falciparum* is commonly defined as a diagnostically-confirmed infection co-presenting with symptoms of disease (most commonly fever) (1). However, this case definition can be problematic in high transmission settings where chronic asymptomatic infections in older children and adults are common (2–6). Individual asymptomatic infections often persist for months without treatment; collectively, these infections serve as a reservoir of infection that perpetuates transmission. Asymptomatically infected individuals who coincidentally seek care for fever of non-malarial etiology may test positive for malaria parasites and be incorrectly diagnosed with clinical malaria, though their *Plasmodium* infection is not the cause of their fever. In high transmission settings, these incorrectly classified febrile episodes may result in overestimation of the incidence of malaria. This presents several problems of practical significance. First, non-malarial febrile illness can be prolonged or worsened when the root cause is not addressed. Second, systematically overestimating the burden of clinical malaria can lead to inappropriate prioritization and use of resources. Third, misclassification of clinical malaria in research trials where it serves as an outcome can compromise power to evaluate new malaria interventions (7). Such trials typically include a threshold parasite density in the malaria clinical case definition – *e.g.*, in Phase-3 trials of the R21 malaria vaccines, the case definition required febrile and parasitologically-confirmed cases to have a parasite density of at least 5000 parasites per microliter of blood (p/µL) (8,9).

The malaria attributable fraction (MAF), *i.e.*, the proportion of clinical malaria cases in which fever is due to infecting parasites, can be used to address these challenges. MAF is estimated through modelled comparison of the parasite density distributions of care-seeking, febrile, individuals with the parasite density distributions of asymptomatically infected individuals (10). The majority of studies estimating MAF use logistic regression models (11–17), but Bayesian latent-class mixture models are more appropriate in high transmission settings where logistic estimates may be biased and imprecise (18,19). MAF Bayesian estimates in high transmission sites in Papua New Guinea in the 1990s and early 2000s ranged from 25-37% and peaked in non-infant children under five (18,19). MAF estimates in lower transmission sites in Kenya and Mozambique ranged from 1-5% (14,20), and MAF was highest in infants (14). Across settings, MAF increased with increasing parasite density and was below 10% in infections with less than 200 p/µL (18,19).

MAF differences across age, transmission intensity, and parasite density levels should be considered when using incidence estimates for national planning, clinical guidelines, and measuring intervention effectiveness. However, MAF estimation to date has had key technical limitations. In particular, available literature calculating MAF largely used parasite densities derived from microscopy rather than qPCR. Parasitemia estimated from microscopy (as compared to qPCR) is less precise and less sensitive for detecting infections with fewer than 100 p/µL; therefore, studies relying only on microscopy have biased parasitemia distributions (21). Additionally, prior MAF estimates were calculated before the modern era of consistent parasitologically confirmed malaria diagnoses, broad test and treat programs, and widespread vector control campaigns. Modern MAF will be higher with more specific case-definitions and its distributions may change in concert with changes in transmission dynamics. Updated MAF estimates generated using modern molecular diagnostics and reliable statistical methods should be incorporated into clinical case definitions to obtain accurate efficacy and effectiveness measures of interventions. This study conducted Bayesian modeling of quantitative PCR (qPCR)-derived parasite densities of asymptomatic and symptomatic infections in a recent Ugandan cohort of all ages followed for up to 3 years to estimate the fraction of fever attributable to malaria.

## Methods

### Study area

The study was embedded in a prospective cohort study (Program for Resistance, Immunology, Surveillance, and Modeling of Malaria (PRISM)) that took place in neighboring Busia and Tororo Districts in eastern Uganda from August 2020 – September 2023. The first and last participant were enrolled on August 10 2020 and February 21 2023, respectively. Both districts have had traditionally high levels of transmission and received long lasting insecticidal treated nets (LLINs) every three years from the national malaria control program starting in 2013. In addition, the malaria program conducted indoor residual spraying (IRS) with insecticide in Tororo beginning in 2014. IRS in Tororo resulted in a dramatic decline in malaria through 2021, followed by a resurgence in 2021 in response to a change to a less effective IRS formulation (22,23). IRS was never conducted in Busia, and transmission there remained high. Based on the intervention history, we identified three zones in the study area: “Busia” zone included individuals living in Busia District, which had the highest transmission intensity; “Tororo, away from border”, included individuals who lived centrally in Tororo District, which had much lower transmission during the time of effective IRS; and “Tororo, near border,” included individuals living in the area of Tororo bordering Busia District, which we expected to have a transmission profile between the first two zones.

### Study design

The PRISM Border Cohort enrolled all permanent residents of 80 randomly sampled households in the three aforementioned zones in Busia and Tororo (24). Cohort study participants were encouraged to come to a dedicated study clinic open seven days per week for all their medical care. In addition, routine visits were conducted every four weeks and included a standardized evaluation and collection of blood samples. The study team sampled all clinical malaria cases and all individuals at monthly routine visits in the cohort to ascertain infection status using qPCR. Study participants found to have a tympanic temperature ≥ 38.0°C or subjective history of fever in the previous 24 hours at the time of visit had a thick blood smear read immediately. If the thick blood smear was positive by light microscopy, the patient was diagnosed with clinical malaria and managed according to national guidelines. Further details on the PRISM Border Cohort, the history of interventions, and the trends in transmission and disease burden are published elsewhere (24).

### Laboratory procedures

Two expert microscopists stained and read blood slides following procedures for parasite density calculations per World Health Organization standards, counting asexual parasites for 200 leukocytes (25). A third microscopist resolved discrepancies as needed. For qPCR, DNA was extracted from 200 µL of whole blood using Qiagen spin columns. Extracted DNA was tested for the presence and quantity of *P. falciparum* DNA using a highly sensitive qPCR assay targeting the multicopy conserved var gene acidic terminal sequence (*var*ATS) with a lower limit of detection of 0.05 p/µL (26). Parasitemia was quantified using serial dilutions of standard curves of parasite genomic DNA from concentrations of 1-10,000 p/µL.

### Ethical approval

The study was approved by the Makerere University School of Medicine Research and Ethics Committee (REF 2019–134) and the University of California, San Francisco Committee on Human Research (257790). All participants or guardians (for minors) completed written informed consent at the time of enrollment.

### Statistical analysis

#### Identification of symptomatic and asymptomatic infections

We used data from routine or febrile visits where an infection was detected by qPCR. We classified infections that were captured during sick visits as “symptomatic” if the individual had a tympanic temperature ≥ 38.0°C or reported fever in the last 24 hours. We classified infections that were captured during enrolment or routine visits as “asymptomatic” if the individual had a tympanic temperature < 38.0°C, did not report fever in the last 24 hours, and did not have a sick visit with malaria diagnosis and treatment within the previous 30 days or following 14 days. Asymptomatic infections with a symptomatic infection within the previous 30 days were excluded to avoid misattribution of parasites from a symptomatic infection to an asymptomatic infection. If any individual had only one asymptomatic infection, and it was during enrollment, we excluded all data from that individual. For any infection to be included, that individual they must have had at least on “symptomatic infection” to ensure population overlap.

#### Malaria attributable fraction model

We implemented a latent class Bayesian model to estimate malaria attributable fraction, lamba (λ). This model was initially described by Vounatsou et al. (10). Parasite densities measured by qPCR in individuals who were febrile (tympanic temperature <u>≥</u> 38.0°C or reported fever) were modeled as a mixture of two distributions i.e. f(x) = (1 ― λ)g_1_(x) + λg_2_(x), where x was the measured parasite density, g_1_(.) and g_2_(.) are the probability density functions of parasite densities of febrile individuals that are not attributable or are attributable to malaria, respectively. Parasite densities in individuals who were asymptomatic were assumed to belong to g_1_(x). We divided our data into eight parasite density levels of p/µL: (0, 0.1], (0.1, 1], (1, 10], (10, 100], (100, 1000], (1000, 10000], (10000, 100000] and (100000, inf), and defined *θ*_i_ = P(x ∈ level_i_| x ∈ P_1_), *ϕ*_i_ = P(x ∈ level_i_|x ∈ P_2_) and p_i_ = P(x ∈ level_i_) = (1 ― λ)θ_i_ +λϕ_i_, where P_1_ and P_2_ were the distribution functions of g_1_ and g_2_ i.e. *λ* = P(x ∈ P_2_). We denoted the number of samples from asymptomatic individuals in level i as m_i_ and the number of samples from febrile individuals in level i as n_i_:

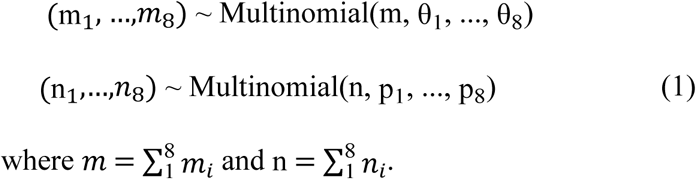

Values of *m*_*i*_ and *n*_*i*_ were obtained by counting the number of asymptomatic and symptomatic visits falling into each density category in our data. It was assumed that a qPCR value in the range of (0, 0.1] was low enough such that the fever of all individuals in that level was considered non-malaria-attributable. It was also assumed that fever was more likely due to malaria with higher qPCR levels, which can be written as:

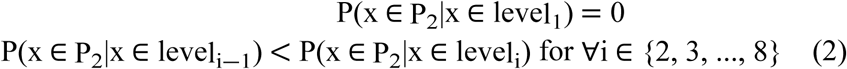

Then *θ*_i_, ϕ_i_ and *λ* can be estimated by maximizing likelihood of (1) given m_i_ and n_i_, under the constraint of (2). It follows that *λ*_i_, the probability that a fever in an individual with parasite density in level *i* is due to malaria, can be calculated as 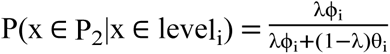. We ran the model stratified by age category (under five years old, five to fifteen years old, and sixteen years old and older), zone, and time period (before and after November 2021). We used the R package afdx to compute the attributable fraction, using the R package rjags to run the models within R. The model ran five parallel chains and checked for convergence visually by adding additional interactions and visually comparing the results and formally using the Gelman-Rubin statistic to test for significant variance across chains (27). Each chain had 5000 burn-in interactions and 10,000 iterations sampling the model parameters to estimate their posterior distributions. We ran a sensitivity analysis with a second set of models that only considered routine or sick visits where an infection was detected by microscopy, though these results focus on visits positive by qPCR.

#### Negative predictive value (NPV) and positive predictive value (PPV) using different parasite thresholds

After estimating *λ*_i_ for all symptomatic infections in the cohort, we calculated the positive predictive value (PPV), negative predictive value (NPV), sensitivity, and specificity of various parasite density thresholds to define clinical cases of malaria. For these calculations we assumed that *λ*_i_ represented the true (gold standard) probability that a fever in an individual with parasite density in level *i was* due to malaria. For each parasite threshold tested, we generated mean estimates and confidence intervals through a bootstrap sampling procedure, where the dataset was resampled 1000 times and, for each observation, *λ*_i_ values were sampled from the posterior corresponding to the specific zone, age-group and parasite density category. We used qPCR parasite thresholds for these curves given the range and precision of the qPCR densities is greater than those of microscopy.

#### Adjusted clinical malaria incidence

We also calculated adjusted clinical malaria incidence for each age category and zone. To do this, we assumed unadjusted clinical malaria incident cases to be febrile infections confirmed positive by microscopy (and thus likely to be diagnosed in a clinical setting). We then weighted each case using the *λ*_i_ estimate specific to the age, zone, qPCR density of the infection. The sum of all cases and all weighted cases were divided by time under observation to estimate uncorrected and MAF-corrected incidence rate. We also calculated unadjusted incidence rates where the case definition included previously used microscopy parasite densities thresholds from 10 to 5000 p/µL.

## Results

The study included 186, 229, and 244 unique participants with at least one follow-up visit and median follow-up time of 683, 1124, and 977 days in each of Busia, Tororo near border, and Tororo away from border zones, respectively (**Table 1**). Study participants in both Busia and Tororo near the border had a median age of 9 and were 53% female. The study population in Tororo away from the border was slightly older and had a higher proportion of female participants. In any given month of the study period, approximately half of the study population was qPCR positive. Average qPCR monthly prevalence was highest in Tororo near the border, followed by Busia, and lowest in Tororo away from the border. Average monthly microscopy prevalence was lower but followed a similar pattern. In all zones, over 95% of individuals were qPCR positive at least once. There were 2195 qPCR positive infections coinciding with fever, which we refer to as “symptomatic infections”, though we use this term as a shorthand for the co-occurrence of fever and infection, rather than necessarily implicating the infection as the cause of the fever.

**Table 1.** Eligible participants included in the final qPCR models.

|  | All regions | Busia | Tororo near border | Tororo away from border |
| --- | --- | --- | --- | --- |
| Number of participants | 659 | 186 | 229 | 244 |
| Number of symptomatic infections | 2195 | 524 | 832 | 839 |
| Number of asymptomatic infections | 6535 | 1557 | 3177 | 1801 |
| Median age (IQR) | 10 (4,24) | 9 (3,22) | 9 (4,22) | 11 (4,24) |
| Proportion female sex (n) | 0.55 (659) | 0.53 (186) | 0.53 (229) | 0.59 (244) |
| Median days of follow-up (IQR) | 977 (536,1125) | 683 (420,1139) | 1124 (733,1131) | 977 (565,979) |
| Microscopy prevalence | 0.24 | 0.21 | 0.31 | 0.19 |
| qPCR prevalence | 0.54 | 0.51 | 0.63 | 0.47 |

Abbreviations: interquartile range (IQR)

As expected, symptomatic infections had higher median parasite densities (1609 p/µL, IQR: 13 - 29626) than asymptomatic infections (7.5 p/µL, IQR: 0.1 - 192), and parasite densities followed predictable age-specific patterns across all zones and time periods (**Figure 1, Table S1**).

**Figure 1.**
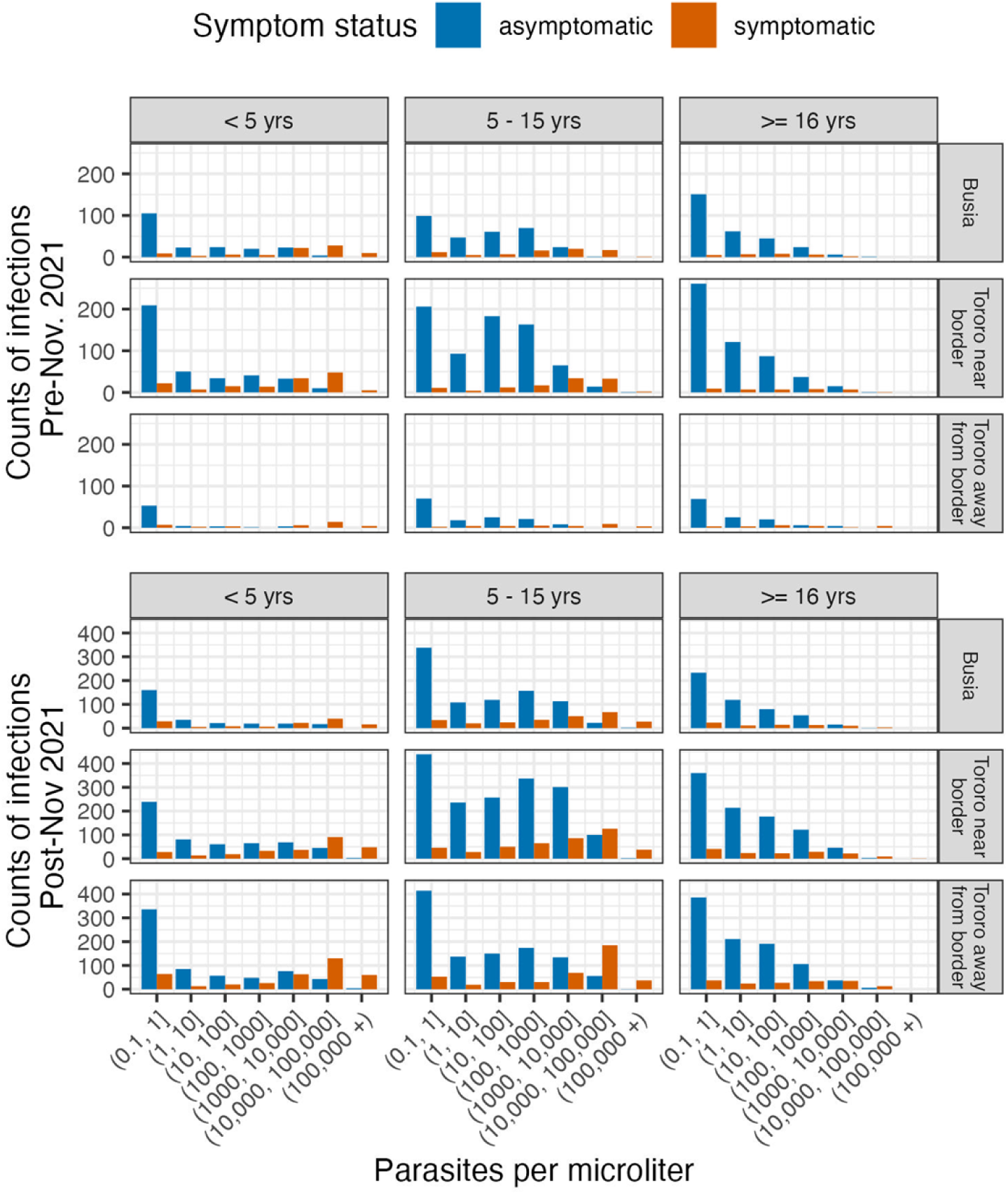
qPCR parasite densities by age, zone, and symptom status in the time periods before and after resurgence of malaria

Asymptomatic infections with parasite density below 1 p/µL were detected at high frequency in all age groups but otherwise appeared at frequencies consistent with increasing immunity with age: in young children, asymptomatic infections were few but occurred at all parasite densities, in children 5-15, asymptomatic infections were common and characterized by moderate and high parasite density levels, and in adults, asymptomatic infection was common and predominantly of low parasite density. Meanwhile, symptomatic infections in children under five and 5 - 15 occurred frequently at all parasite densities and most frequently at high densities, whereas symptomatic infections in adults were less common and parasite densities were rarely high.

### Malaria attributable fraction varied by transmission intensity, age, and time

We estimated that only 58% of fevers with parasitemia detectable by qPCR were attributable to malaria (MAF), with substantial variation by zone, age group, and study time (**Table 2**). MAF was generally lower in zones where prevalence was higher. Across all strata of zones and time, MAF was highest in under 5-year-olds (72% overall), lower in 5-15 year-olds (56%) and lowest in adults over 16 years old (45%). Across most strata of age and zones, MAF was higher in the period before the resurgence (August 2020-November 2021) than after, with the greatest difference (13 and 21 percentage point drop) in Tororo zones where the resurgence occurred.

**Table 2.** Malaria attributable fraction by zone, age, and time-period. Prior to November 2021, malaria was controlled in Tororo District by effective indoor residual spraying (IRS). Following a change in insecticide used in IRS, malaria resurged in Tororo after November 2021.

| **All zones** |  |  |  |
| --- | --- | --- | --- |
|  | Overall (95% CI) | Pre Nov 2021 (95% CI) | Post Nov 2021 (95% CI) |
| All-age | 0.577 (0.439, 0.731) | 0.688 (0.453, 0.880) | 0.557 (0.403, 0.727) |
| Under 5 year-olds | 0.719 (0.599, 0.833) | 0.768 (0.596, 0.916) | 0.692 (0.548, 0.828) |
| 5-15 year-olds | 0.556 (0.450, 0.691) | 0.707 (0.482, 0.864) | 0.552 (0.428, 0.697) |
| 16+ year-olds | 0.445 (0.250, 0.666) | 0.559 (0.239, 0.852) | 0.415 (0.217, 0.651) |
| **Busia (Prevalence overall: 51%, pre-resurgence: 46%, post-resurgence: 54%)** |  |  |  |
| All-age | 0.558 (0.394, 0.741) | 0.611 (0.384, 0.837) | 0.558 (0.372, 0.765) |
| Under 5 year-olds | 0.671 (0.554, 0.790) | 0.776 (0.602, 0.913) | 0.610 (0.476, 0.758) |
| 5-15 year-olds | 0.568 (0.422, 0.737) | 0.610 (0.416, 0.802) | 0.607 (0.422, 0.799) |
| 16+ year-olds | 0.415 (0.174, 0.689) | 0.380 (0.033, 0.778) | 0.442 (0.193, 0.733) |
| **Tororo, near border (Prevalence overall: 63%, pre-resurgence: 63%, post-resurgence: 70%)** |  |  |  |
|  | Overall | Pre Nov 2021 | Post Nov 2021 |
| All-age | 0.536 (0.416, 0.672) | 0.630 (0.434, 0.830) | 0.504 (0.358, 0.662) |
| Under 5 year-olds | 0.769 (0.643, 0.875) | 0.729 (0.545, 0.886) | 0.763 (0.598, 0.893) |
| 5-15 year-olds | 0.475 (0.377, 0.601) | 0.606 (0.450, 0.792) | 0.437 (0.323, 0.589) |
| 16+ year-olds | 0.364 (0.228, 0.541) | 0.543 (0.284, 0.809) | 0.311 (0.154, 0.503) |

| **Tororo, away from border (Prevalence overall: 47%, pre-resurgence: 18%, post-resurgence: 58%)** |  |  |  |
| --- | --- | --- | --- |
|  | Overall | Pre Nov 2021 | Post Nov 2021 |
| All-age | 0.636 (0.507, 0.779) | 0.824 (0.543, 0.974) | 0.608 (0.480, 0.754) |
| Under 5 year-olds | 0.717 (0.600, 0.835) | 0.800 (0.641, 0.949) | 0.703 (0.570, 0.834) |
| 5-15 year-olds | 0.625 (0.550, 0.733) | 0.905 (0.581, 0.998) | 0.612 (0.541, 0.703) |
| 16+ year-olds | 0.556 (0.349, 0.766) | 0.753 (0.401, 0.969) | 0.492 (0.304, 0.718) |

### Malaria attributable fraction was substantial even at low parasite densities

We estimated MAF within parasite density strata of *i* (where *i* is 0.1 - 1 p/µL, 1-10 p/µL, etc) as *λ*_i_. While estimates for *λ*_i_ were consistent with prior evidence that a greater proportion of febrile illnesses are attributable to the malaria parasite at higher parasite densities, there are a few surprising observations across the population and noteworthy group-level deviations from population trends (**Figure 2, Table S2**). First, while lower confidence bounds of *λ*_i_ in infections with a parasite density < 1 p/µL were low, they were not zero, and the point estimate was often well above zero, suggesting infections with very low densities can cause or at least contribute to disease in individuals of any age (**Table S2**). Second, *λ*_i_ values were often above 30% in infections of parasite density 10 - 100 p/µL (a range in which standard diagnostics are only moderately sensitive) (28) and above 50% in infections of parasite densities 100 - 1000 p/µL. Lastly, *λ*_i_ values were well above 50% in infections of 1000 - 10000 p/µL: the average MAF of this density strata, across all ages and zones was 76%.

**Figure 2.**
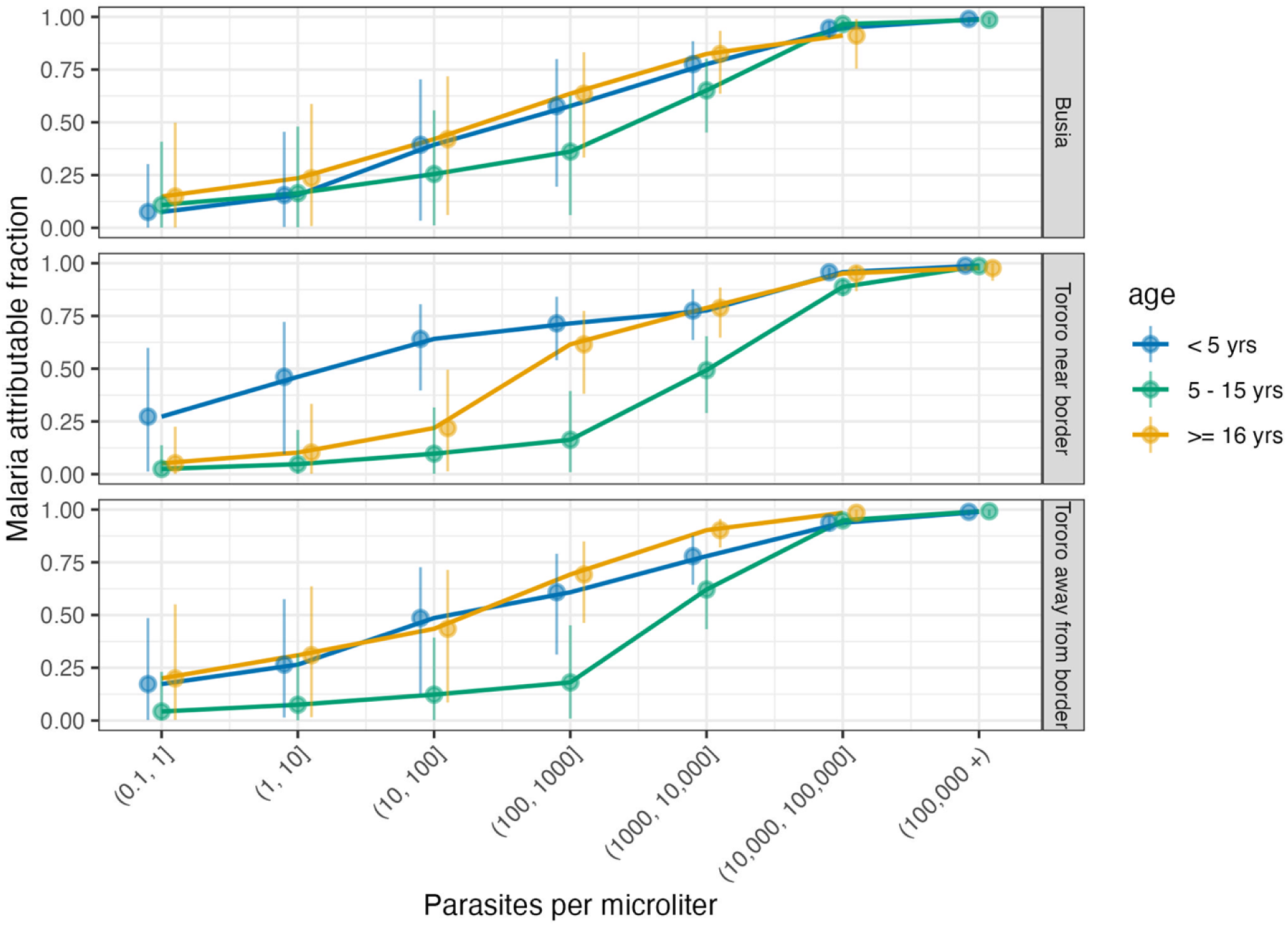
Full study period malaria attributable fraction by zone, age, and parasite density

Site-level age-trends in *λ*_i_ reflect site transmission, transmission history, and surprising differences between adults and children 5-15 years old. In Busia District (**Figure 2, top panel**), where transmission has been consistently high in the setting of no IRS, there was little differentiation in *λ*_i_ by age group. In contrast, in Tororo away from the border (**Figure 2, bottom panel**), where transmission was low then rebounded in 2021, overall prevalence is similar to Busia but *λ*_i_ is higher, and this difference was largely driven by the adult population; adults had higher *λ*_i_ than 5-15 year olds, particularly in infections of low to moderate densities 100 - 1000 p/µL. In Tororo near the border (**Figure 2, middle panel**), where individuals had relatively more parasite exposure compared to those away from the border prior to resurgence, adult *λ*_i_ was similar to 5-15 year-olds at very low-densities, but as parasite densities increased, adult *λ*_i_ rose sooner than in 5-15 year olds. While *λ*_i_ followed expected trends with respect to parasite density for all age groups, *λ*_i_ stratified by parasite density being higher in adults than in 5-15 year olds was surprising: this appeared to be in part driven by higher *λ*_i_ in female adults at all sites (**Figure S2**).

### Age-specific parasite thresholds in case definitions may be needed to maximize negative predictive value (NPV) and positive predictive value (PPV)

Malarial and non malarial attributable cases by parasite density (**Figure 3, Panel A**) informed calculations of positive predictive value (the proportion of febrile infections above a given parasite threshold that are attributable to the malaria parasite) and negative predictive value (the proportion of febrile infections below a given parasite density that are not attributable to the malaria parasite), which varied by age group (**Figure 3, Panel B**). In children under five years old both the PPV and NPV were above 80% even at very low parasite thresholds. In contrast, in 5-15 year olds and in adults, there was a greater tradeoff between PPV and NPV. In 5-15 year olds a parasite threshold of 1000 p/µL was required to achieve a PPV of 80%, and this maintained a NPV of 80%. In adults, achieving a PPV of 80% required a parasite threshold of at 200 p/µL and came at the expense of a lower NPV. NPV and PPV varied across the three relatively high transmission zones of this study, but the general age trend was consistent.

**Figure 3.**
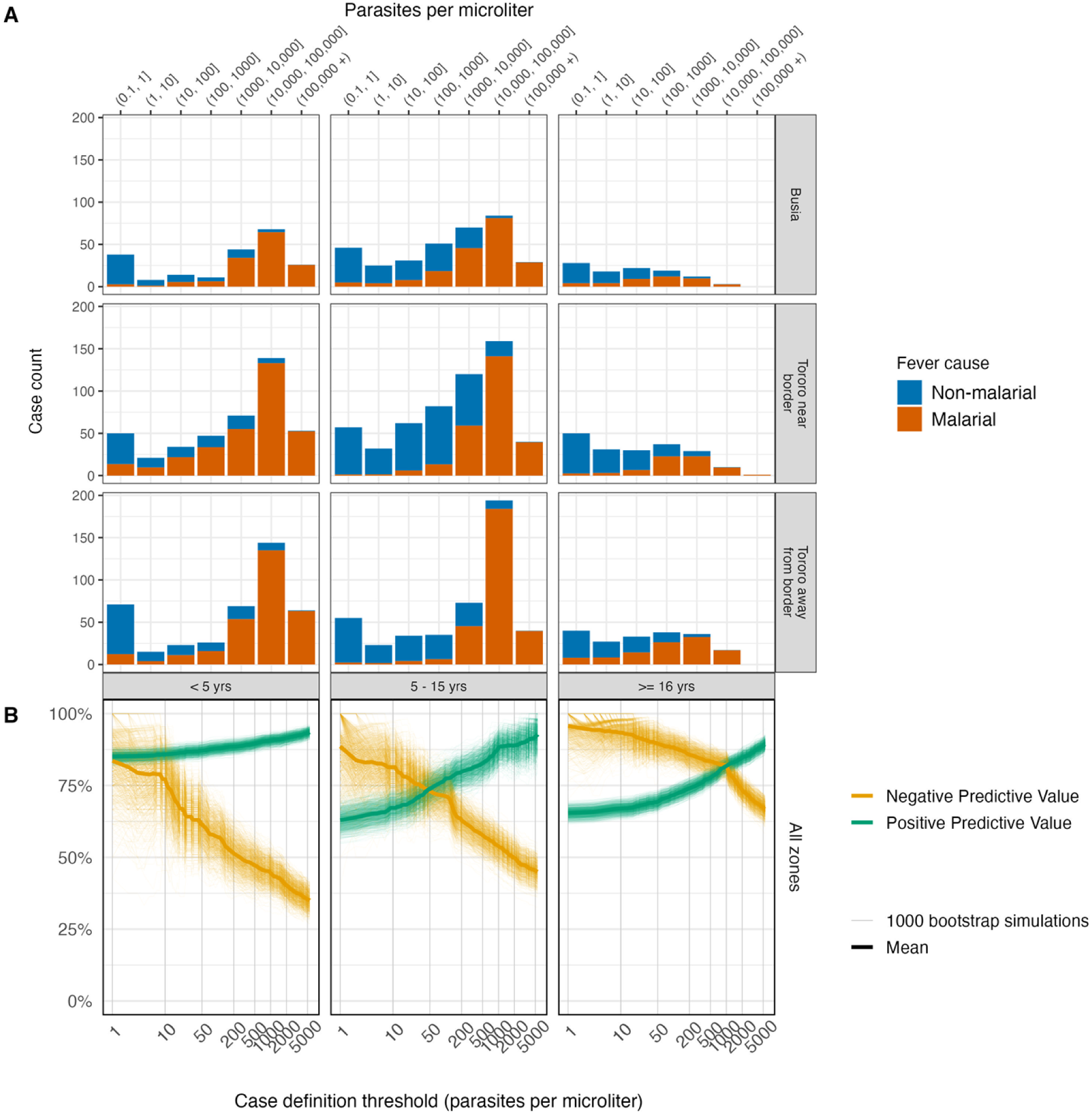
Panel A. Number of febrile, microscopy-positive cases classified as attributable or not to malaria per the qPCR model by site, age group, and parasite density. Panel B. Negative and positive predictive value of defining clinical malaria cases using different qPCR parasite thresholds, using *λ*_i_ estimates as the gold standard.

Accordingly, the NPV and PPV of given parasite threshold case definitions resulted in different absolute case counts. Of the 2226 total febrile microscopy-positive infections in the cohort, 1256 were above 5000 p/µL and met the clinical case outcome definition in vaccine trials, but 1708 were estimated to be attributable malaria. The trial definition with a 5000 p/µL missed 452 cases or 26% of malaria attributable cases in this context. Redefining these thresholds, using age-specific cutoffs that maximize the NPV and PPV would recover many missed cases: even a non-age-specific lower threshold of 1000 p/µL would regain 339 or 75% of missed cases.

### “MAF-corrected” incidence estimates suggest that parasite thresholds used in clinical trials may severely underestimate incidence in many settings

Uncorrected incidence rate overestimated MAF-corrected incidence rate by 30% (1744 vs 1338 cases per 1000 person-years (**Figure 4**), but the incidence rate of infections with at least 5000 p/µL (the case definition for clinical malaria in vaccine trials) underestimated MAF-corrected incidence rate by 20% (1073 vs 1338 cases per 1000 person-years). The 5000 p/µL case definition underestimated MAF-corrected incidence-rate in all zone-specific age-groups (underestimation was 37% on average and ranged from 2% to 66%) except 5-15 year olds in Tororo near the border where the number of incident cases were nearly identical (**Table S3**). On average, the 5000 p/µL case definition underestimated incidence by 18% in children under five (the target population of many of these studies), by 7% in 5-15 year olds, and by 70% in adults but there was nuance to these patterns by zone. For example, in both Tororo zones, the 5000 p/µL case definition underestimated MAF-corrected incidence by only 2% and by 0% in 5-15 year olds, suggesting 5000 p/µL may be an appropriate cutoff for clinical cases in 5-15 year olds in these areas. However, all other zone- and age-specific MAF-corrected incidence rates, including in 5-15 year olds, were substantially underestimated by the 5000 p/µL case definition.

**Figure 4.**
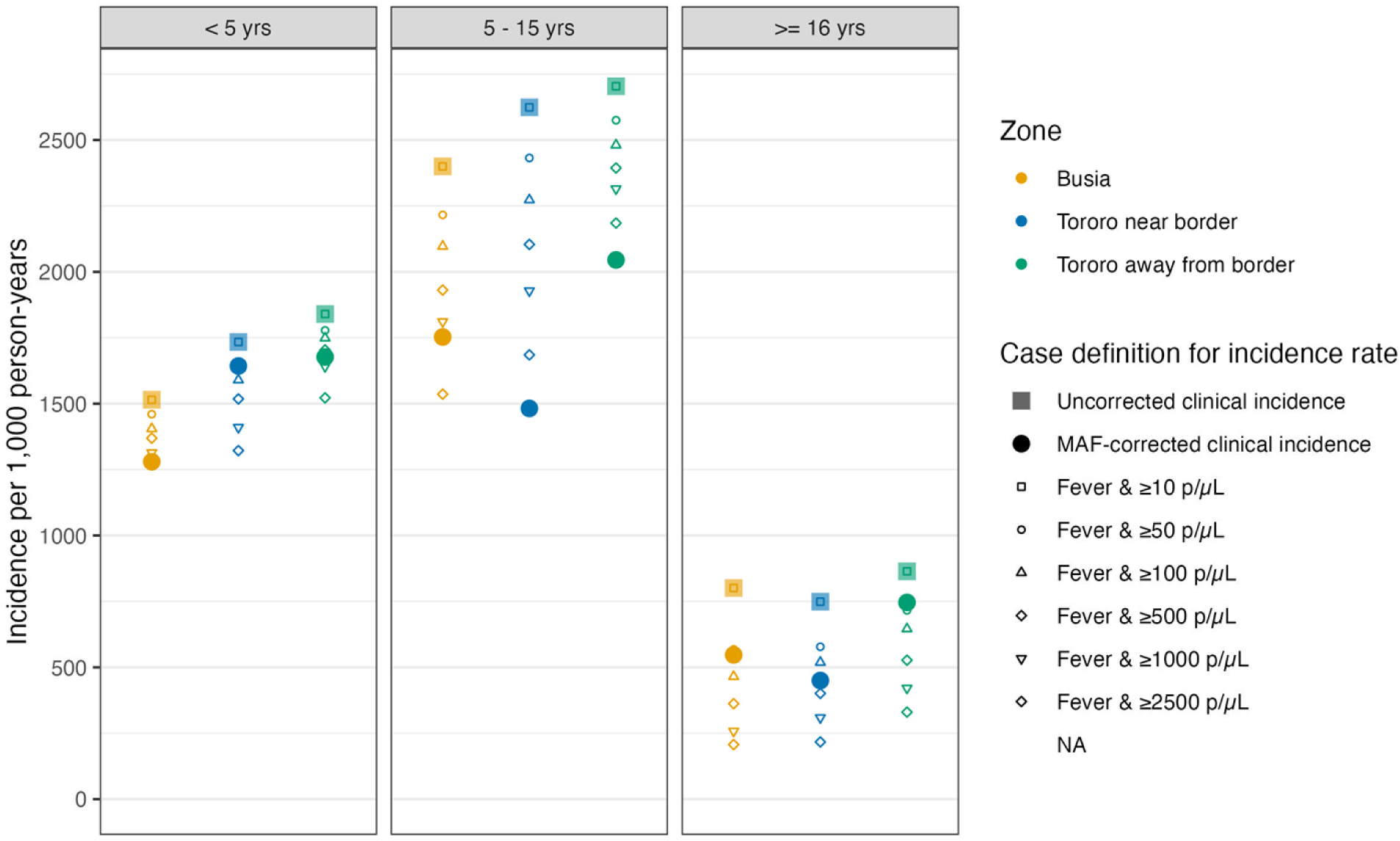
Incidence rates by zone and age group. Uncorrected clinical incidence was the incidence of febrile, microscopy-confirmed cases diagnosed in the clinic. To obtain MAF corrected incidence, microscopy-confirmed cases counts multiplied by the qPCR-density, age, and zone-specific malaria *λ*_*i*_ generated from the model using qPCR data. Clinical incidence ≥ 5000 p/µL is the incidence of febrile, microscopy-confirmed cases, diagnosed in the clinic with parasite densities at or above 5000 p/µL used in vaccine trials and other thresholds are also shown.

### Estimates of MAF using microscopy followed similar trends

Apart from the expected absence of very low density microscopy-detected infections, microscopy and qPCR parasite density distributions across zones and age groups were similar (**Table S4, Figure S4**). MAF calculated using microscopy data demonstrated similar trends across zone, age, and period as the corresponding qPCR estimates, including MAF values well above zero for infections with low parasite density (**Table S6, Figure S5**).

## Discussion

This study leveraged a Bayesian approach to estimate malaria attributable fraction using parasite densities of asymptomatic and symptomatic infections sampled from a cohort in eastern Uganda. Its key finding, that MAF was nearly 50% in low density infections, may shift the current understanding of malaria attributable disease in these populations. Other studies, limited by their microscopy-estimated densities, could not evaluate MAF at low parasite density levels and did not report as high MAF at moderate density levels (11,15). The large and well-described reservoir of asymptomatic, moderate-density infections in high intensity settings (4,6) has motivated the use of a pyrogenic threshold of 5000 p/µL in the primary clinical case definition in RTS,S and R21 malaria vaccine trials (8,9,26,29–31). However, our data suggests that, while highly specific, this threshold is most applicable to older children and misclassifies a high proportion of infections in other age groups. The application of such a threshold to these data (across all age groups) would misclassify 26% of malaria attributable symptomatic infections as not attributable to malaria. This misclassification would be differential according to the transmission intensity experienced by a population and the age strata within a population; in certain age-groups this threshold would misclassify up to 76% of infections, greatly diminishing power, and possibly biasing estimation of age and site-level effects. Using a single dichotomized parasite density threshold is overly simplistic in studies estimate intervention effectiveness across varied transmission settings with populations with differing immunological profiles. This be particularly problematic when the intervention targets young children, for whom a lower threshold is indicated. Alternatively, where resources allow, studies could collect preliminary data to estimate MAF in their study areas and use age and site-specific MAFs to set parasite thresholds that maximize the number of cases while maintaining a minimum acceptable negative predictive value. An alternative method used by vaccine trials has been to conduct sensitivity analyses using multiple parasite density thresholds (9).

Another distinct finding in this study was that *λ*_i_ in adults with low to moderate levels of parasite densities was higher and more similar to *λ*_i_ in young children, whereas *λ*_i_ in children 5-15 years of age remains low, even as we expect adults and older children to have similar immunity profiles. The acquisition of anti-parasite and anti-disease immunity over the lifecourse is consistent with the observed *λ*_i_ patterns in children. In young children, infections are typically symptomatic due to low anti-parasite and anti-disease immunity; consistent with this, we observed high parasite densities in symptomatic infections and very low parasite density in asymptomatic infections (32–34). Older children appear to maintain lower anti-parasite immunity, but develop high anti-disease immunity (35,36). This is consistent with our observations of parasite density (**Figure 2**) in 5-15 year-olds where high parasite densities in symptomatic infections reflect low anti-parasite immunity, while relatively high parasite densities in asymptomatics reflect high anti-disease immunity. The elevated *λ*_i_ in adults at low-to-moderate densities may be explained by a combination of robust anti-parasite and anti-disease immunity in this age group (37). Adults have consistently lower parasite densities and incidence of malaria compared to children. However, when adults do develop parasitemia the malaria attributable fraction may be higher that 5-15 year-olds because of a lower risk of non-malarial causes of fever. Alternative explanations for the higher *λ*_i_ in adults may be biological or may be an artifact of other phenomena that inform *λ*_i_. Higher adult *λ*_i_ may be biologically meaningful if female participants, when pregnant, have enhanced susceptibility to clinical malaria, or other underlying immunological differences independent of pregnancy; the adult study population did have a disproportionate number of female participants and a higher MAF in females as compared to males. This sex difference could also be explained through differential care seeking – female adults more often accompany children to the study clinic and may have more opportunities to test than male adults. Differential incidence of other causes of fever and differential care seeking could also explain the differences in *λ*_i_ between adults and older children at low to moderate density. This study’s data did not provide substantial evidence to support or refute these hypotheses, and the relatively high uncertainty (wide confidence intervals) of the age and density-stratified *λ*_i_ estimates limit our conclusions regarding these trends.

Notable differences in the study designs, settings, and estimation methods may explain the dramatically higher overall MAF in this study (58% overall) as compared to previous Bayesian models that produced MAF estimates ranging from 25 to 37% (19,20,26). While this study estimated MAF with qPCR rather than microscopy densities, a more important difference was the use of parasitological confirmation (via RDT or microscopy) in the case definition. Studies based on data from the 1990s - 2000s calculated MAF in the context of clinical, but not diagnostically confirmed, malaria, driving the denominator of MAF up, and MAF itself, down. There are also true contextual and population differences limiting the comparability of MAF across this and older studies. Broad vector control was not standard in the ‘90s and early ‘00s, while household IRS coverage in Tororo during the study period was 80 - 100%. Participants in this cohort were sensitized to present at the study clinic for all care, and prompt and effective antimalarial therapy was provided free of charge. For these reasons, severe malaria was rarely diagnosed. Cohort members may have been more likely to seek care, present at earlier stages of illness, or present with non-malarial fever than care-seeking individuals in more “real world” settings, which could inflate the denominator of MAF and bias MAF downwards (23). Lastly, differences in malaria attributable fraction could be due to differing rates of other fever-causing illnesses that cause asymptomatically malaria-infected individuals to seek care. Coverage of vaccination against preventable fever-causing childhood illnesses, while still relatively low in Africa, has increased drastically globally over the last three decades (38,39). Our study period also coincided with the emergence and spread of SARS-CoV-2, and serological testing of this study population revealed 96% had been infected by either the Delta or Omicron variant by the end of 2022, likely increasing prevalence of febrile disease and affecting care-seeking behaviors (39,40). This highlights one of the inherent limitations of malaria attributable fraction - it is a function of multiple phenomena including the distributions of symptomatic and asymptomatic infections across parasite densities, the prevalence of other non-malaria infections that lead to care seeking in parasite positive individuals, individual-level behavior, and exposure experienced based on their environment. Further, the applications of MAF are context and parasite-level dependent. While we’ve discussed the benefits of this study’s use of qPCR, we must also consider when MAF estimated for these lowest density categories is useful (in the context of laboratory and clinical trials) and when these lowest density categories may be irrelevant (in a clinical setting when diagnostic methods would not detect them). The heterogeneity in MAF even within this study tells us we must be cautious in making assumptions about MAF outside of the age, density, and transmission intensity in which they are estimated.

A strength of this study is the use of Bayesian latent class models, which are less prone to bias in high transmission areas, better leverage data to yield more precise estimates, and have better statistical fit than logistic and binary models (10,41). Routine sampling every 4 weeks (active surveillance) and care provided 7 days/week for all illnesses (passive surveillance) provided near-complete distributions of parasitemia in symptomatic and asymptomatic infections in the same population. Thus, the distributional comparison of symptomatic and asymptomatic paristemias minimizes biases introduced from imperfect geographic or temporal overlap. Embedding these models in a cohort is also beneficial as we have three continuous years of data, rather than repeated cross-sectional data at limited time points. This is a distinct advantage of this study over other MAF studies, where one or two cross-sectional surveys were used to construct the asymptomatic parasite distributions.

Malaria attributable fraction can be informative to clinical practice and resource management through corrected incidence rates, and for epidemiologic study design of interventions trials and evaluations. It is not surprising that MAF across levels of parasite density, age, and transmission environment is highly variable: this is driven by environmental (endemicity and exposure) and individual (age, immunity) factors (Smith, 1994, Armstrong Schellenberg 1994), many of which are not measurable. Here, we estimated MAF with respect to factors that are measurable (age, site, parasite density) and demonstrated the degree to which traditional definitions may misclassify clinical incidence of malaria, complicating planning and threatening the validity of scientific results. These findings indicate a more nuanced approach to estimating clinical incidence of malaria is needed; these nuances could be as complex as estimating MAF in a given study population or the use of more crude thresholds to adjust incidence by age and transmission level.

## Data Availability

Data from the cohort study and health facility-based surveillance will be made available through an established open-access clinical epidemiology database resource, ClinEpiDB.

## Acknowledgments

We thank all the study team members for successfully conducting the PRISM studies over the years and the Tororo and Busia district administrations for their support. We are grateful to the study participants who participated in this study and their families.

